# The Role of GLP-1 Receptor Agonists in Managing Cancer Therapy-Related Cardiac Dysfunction

**DOI:** 10.1101/2025.01.02.25319923

**Authors:** Aravinthan Vignarajah, San Kim, Moath Albliwi, Hyunjun Max Ahn, Aleksandar Izda, Faris Naffa, Nishanthi Vigneswaramoorthy, Shimoli Barot, Gautam Shah

## Abstract

**Background:** Anti-cancer therapies currently available can lead to cancer therapy-related cardiac dysfunction (CTRCD). Glucagon-like peptide-1 receptor agonists (GLP-1 RAs) have been shown to exhibit cardioprotective effects but its impact on CTRCD has not been evaluated.

**Objectives:** This study evaluates the impact of GLP-1 RAs on all-cause mortality, hospitalization, and heart failure exacerbation in patients with CTRCD.

**Methods:** This is a retrospective cohort analysis using the TriNetX research network. Patients aged ≥18 years with a history of cancer who received antineoplastic therapies and subsequently developed cancer therapy-related cardiac dysfunction (CTRCD) between January 1, 2012, and January 1, 2023, were included. Patients receiving guideline-directed medical therapy (GDMT) for heart failure were divided into two groups: those using GLP-1 RAs and those not using them. Propensity score matching (1:1) was applied based on demographics, comorbidities, and medications, resulting in a matched cohort of 1,223 patients. Outcomes over a 1-year follow-up were measured.

**Results:** The study cohort found 12,410 patients with CTRCD of which 1,223 had received GLP-1 RA treatment (mean age: 66.4 years; 47.3% female; 65.9% White). Patients receiving GLP-1 RAs in addition to GDMT had a significantly lower risk of acute heart failure exacerbations, all-cause mortality, and all-cause hospitalization (OR: 0.566 [95% CI: 0.48-0.668]; P < 0.001). A clinically significant reduction in atrial fibrillation/flutter and ventricular tachycardia was observed in the GLP-1 RA groups, although did not reach statistical significance.

**Conclusion:** In patients with CTRCD, adding GLP-1 RA therapy to GDMT significantly improves mortality and heart failure outcomes over 1 year.

## INTRODUCTION

Cancer therapeutics are evolving at an unprecedented rate and many new anti-cancer treatments are introduced every year. With this, the number of new and existing anti-cancer agents causing cancer therapy-related cardiac dysfunction (CTRCD) is also increasing (1). About 10-19% of patients undergoing conventional cancer therapies develop CTRCD, defined as a reduction in left ventricular ejection fraction (LVEF) of more than 10% or to a value below 53% due to cancer therapy (2). The mortality risk from heart failure (HF) related to chemotherapy is 3.5 times higher than that of idiopathic cardiomyopathy and the long-term risk of mortality from cardiovascular complications surpasses the risk of tumor recurrence (3).

Glucagon-like peptide-1 receptor agonists (GLP-1 RAs), initially introduced as anti-diabetic medications, have demonstrated cardioprotective effects far beyond their glycemic control (4). Several randomized controlled trials have established the role of GLP-1 RAs in improving cardiovascular outcomes in patients with T2DM making them the standard of care for heart failure patients (5).

Recent animal studies demonstrated a cardioprotective effect of GLP-1 RA in rats treated with anti-cancer therapies, primarily by its antioxidant and anti-inflammatory properties and by improving cardiac biomarkers and cardiac function (6). Despite these favorable findings, many clinical trials proving the cardioprotective effects of GLP-1 RAs have excluded patients with anti-cancer treatment (5). Due to this, the application of GLP-1 RAs in CTRCD patients remains unvalidated. This study seeks to investigate the impact of GLP-1 RAs on cardiovascular outcomes in patients who have developed CTRCD.

## METHODS

### Data Source

We used TriNetX, a global federated health research network providing access to statistics on electronic medical records (diagnoses, procedures, medications, laboratory values, genomic information) from approximately 134 million patients in 97 large Healthcare Organizations, predominantly in the United States. As a federated network, TriNetX received a waiver from Western IRB since only aggregated counts, statistical summaries of de-identified information, but no protected health information is received, and no study-specific activities are performed in retrospective analyses.

### Study Population and Design

The study population consisted of patients aged 18 years and older with a history of cancer who had undergone treatment with cardiotoxic antineoplastic therapies and subsequently developed cancer therapy-related cardiac dysfunction (CTRCD). The study included patients treated between January 1, 2012, and January 1, 2023. The antineoplastic agents evaluated for their cardiotoxic effects included anthracyclines, alkylating agents, antimetabolites, monoclonal antibodies, small-molecule tyrosine kinase inhibitors, and proteasome inhibitors. These medications were selected based on their cardiotoxic properties reported in prior studies (7). Patients with a history of acute coronary syndromes or those who had undergone coronary artery bypass grafting (CABG) or percutaneous coronary intervention (PCI) were excluded from the study to exclude the potential confounding effect of ischemia-induced cardiac dysfunction. Cancer history and heart failure were identified using ICD-10 codes. All participants received optimal guideline-directed medical therapy (GDMT), including angiotensin-converting enzyme inhibitors, angiotensin receptor blockers, or angiotensin receptor-neprilysin inhibitors, beta-blockers, and mineralocorticoid receptor antagonists in differing combinations. The study population was divided into two groups: those who received GLP-1 receptor agonists (GLP-1 RA) and those who did not.

### Study Outcomes

The study analyzed primary and secondary outcomes over a 1-year follow-up period. The primary outcomes included HF exacerbations (identified by ICD-10 codes or the need for intravenous loop diuretics), all-cause hospitalization or emergency department visits, and all-cause mortality. Secondary outcomes included all-cause hospitalizations or emergency department (ED) visits, all-cause mortality, heart failure exacerbation, atrial fibrillation/flutter, and ventricular tachycardia. Gastroenteritis was analyzed as a falsification outcome.

### Statistical Analysis

The patient cohort was categorized into two groups based on whether they were prescribed GLP-1 receptor agonists (GLP-1 RA). Continuous variables are presented as mean ± standard deviation (SD) and compared using independent-sample t-tests, while categorical variables are expressed as n (%) and analyzed via the chi-square test. To address baseline differences between the cohorts, 1:1 propensity-score matching (PSM) was applied using a built-in algorithm that utilizes a greedy nearest-neighbor method with a caliper of 0.1 pooled SDs. Variables with a P-value of >0.05 were considered well-matched. All variables in **Table 1** were analyzed based on the outcomes using the multivariable logistic regression model. Survival analysis was conducted by plotting Kaplan-Meier curves and comparing the two cohorts with log-rank tests. Statistical significance was defined by a two-sided P value of <0.05. All statistical analyses were performed using the TriNetX online platform, with R for statistical computing.

**Table 1:**
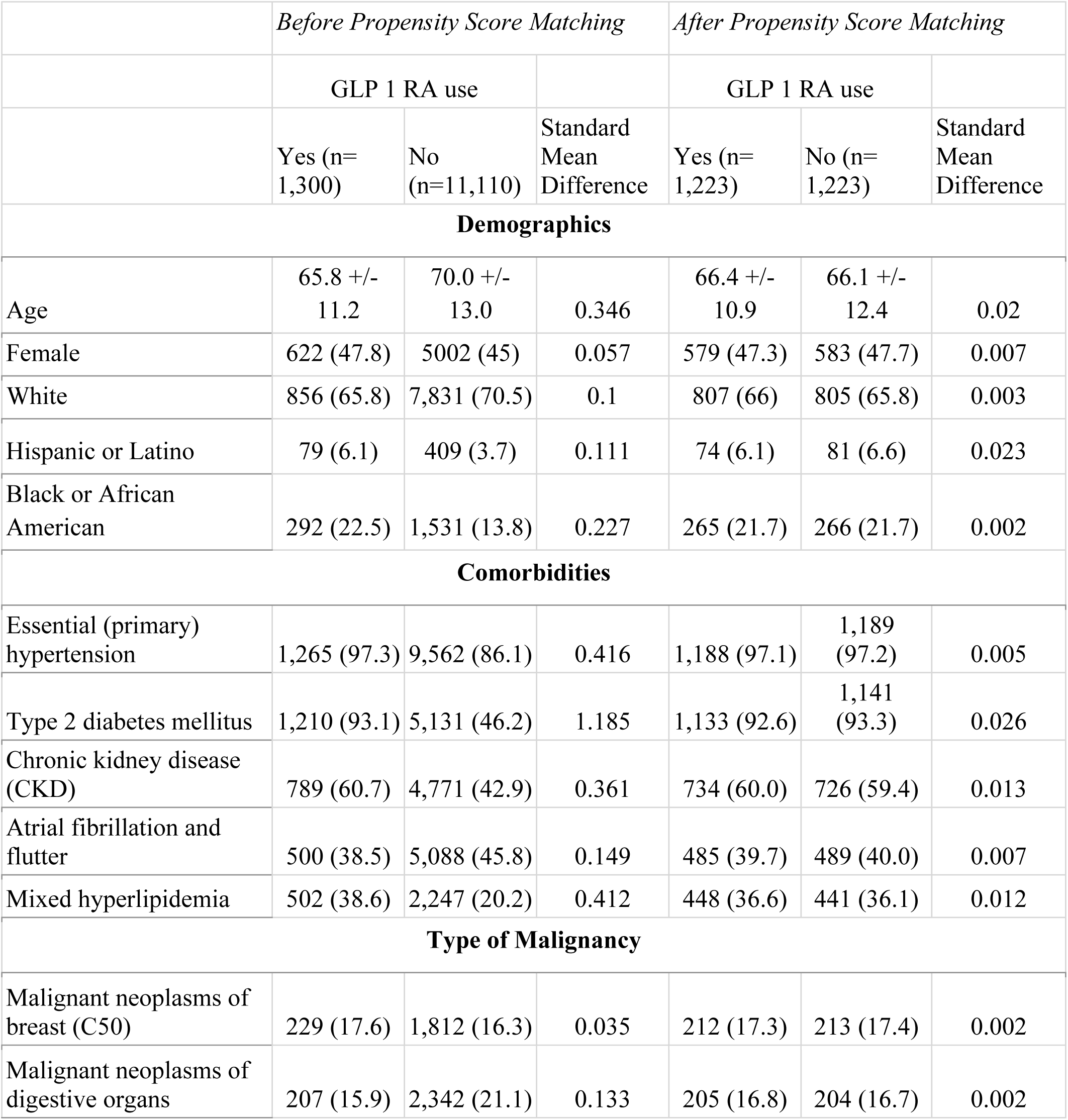
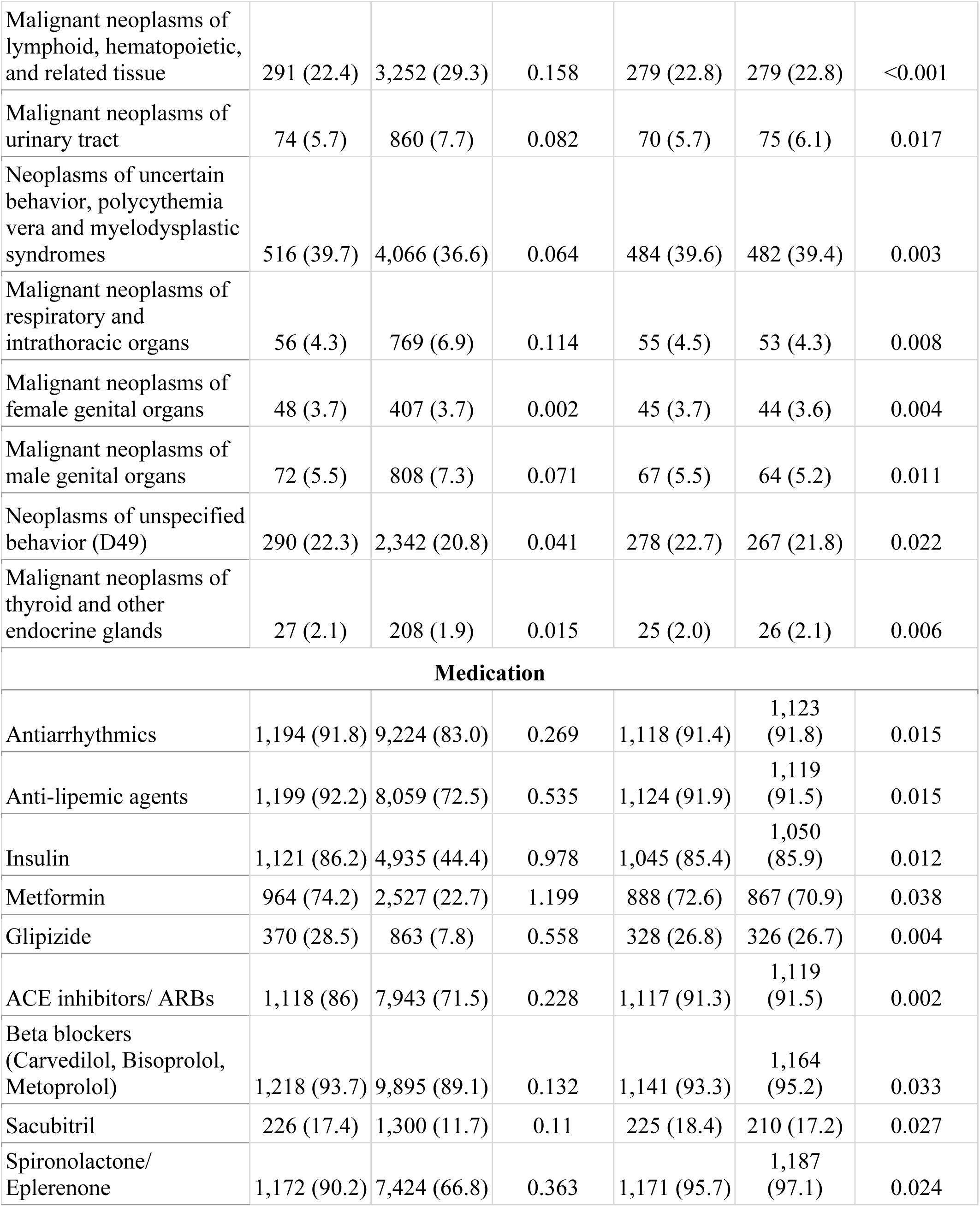
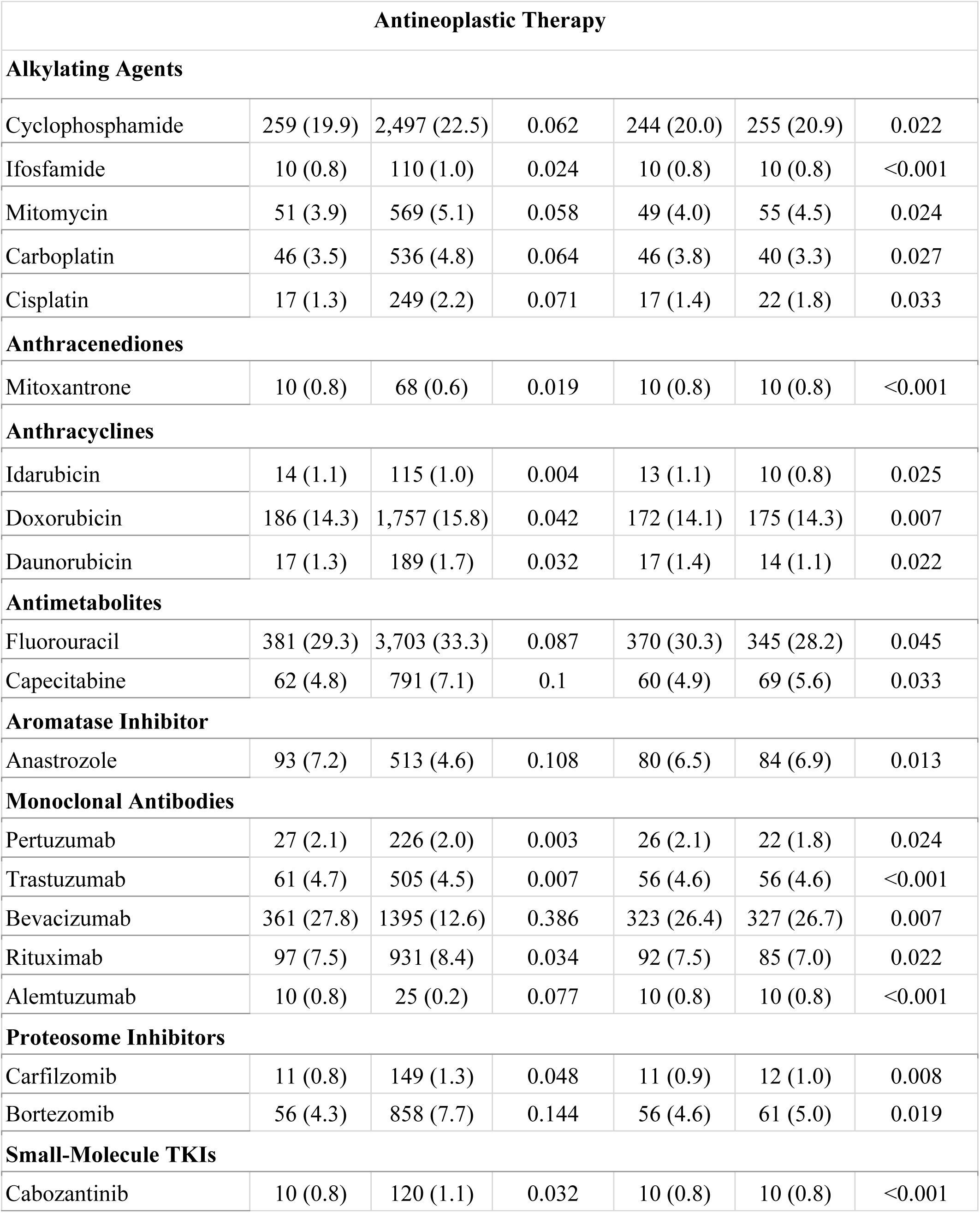
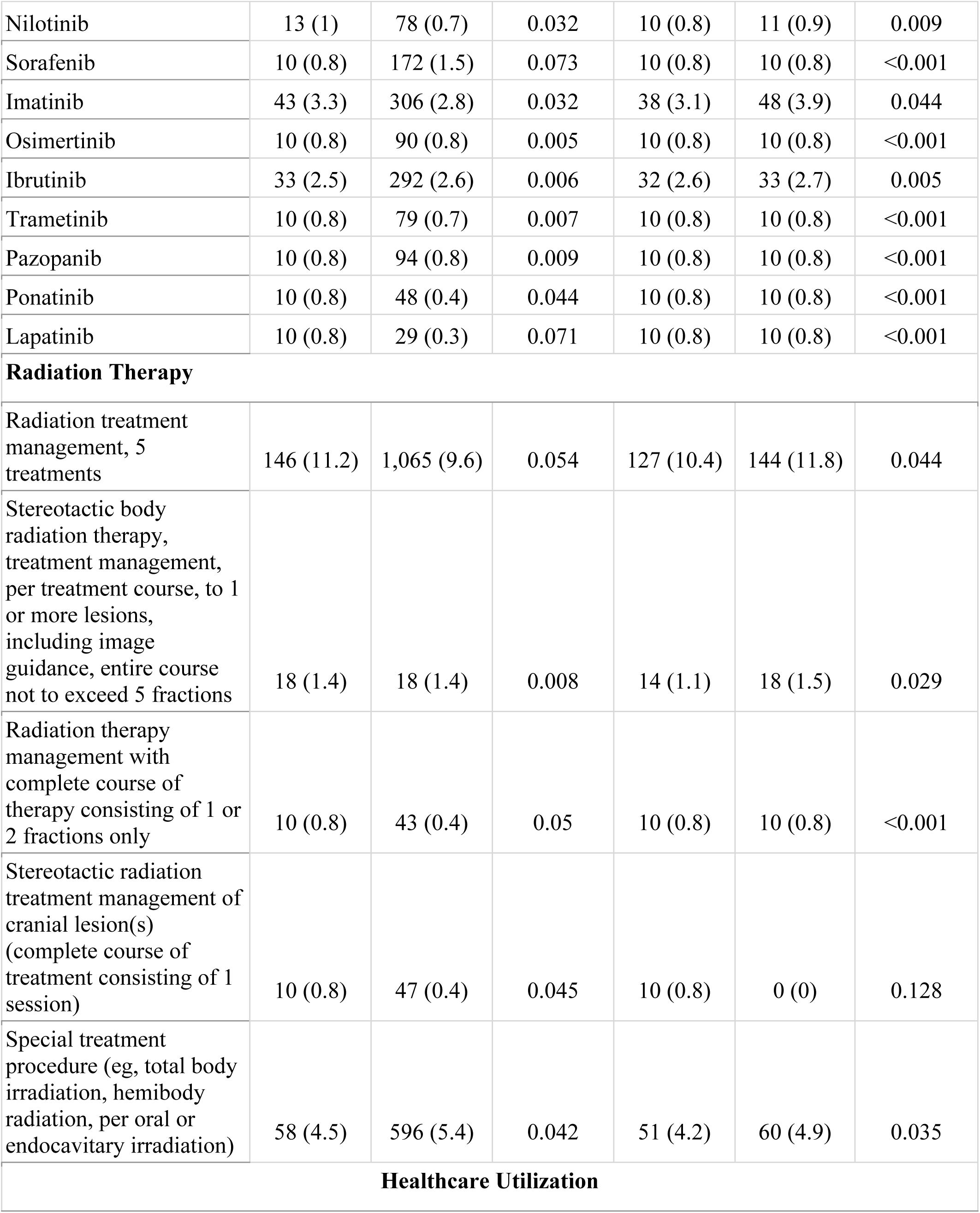
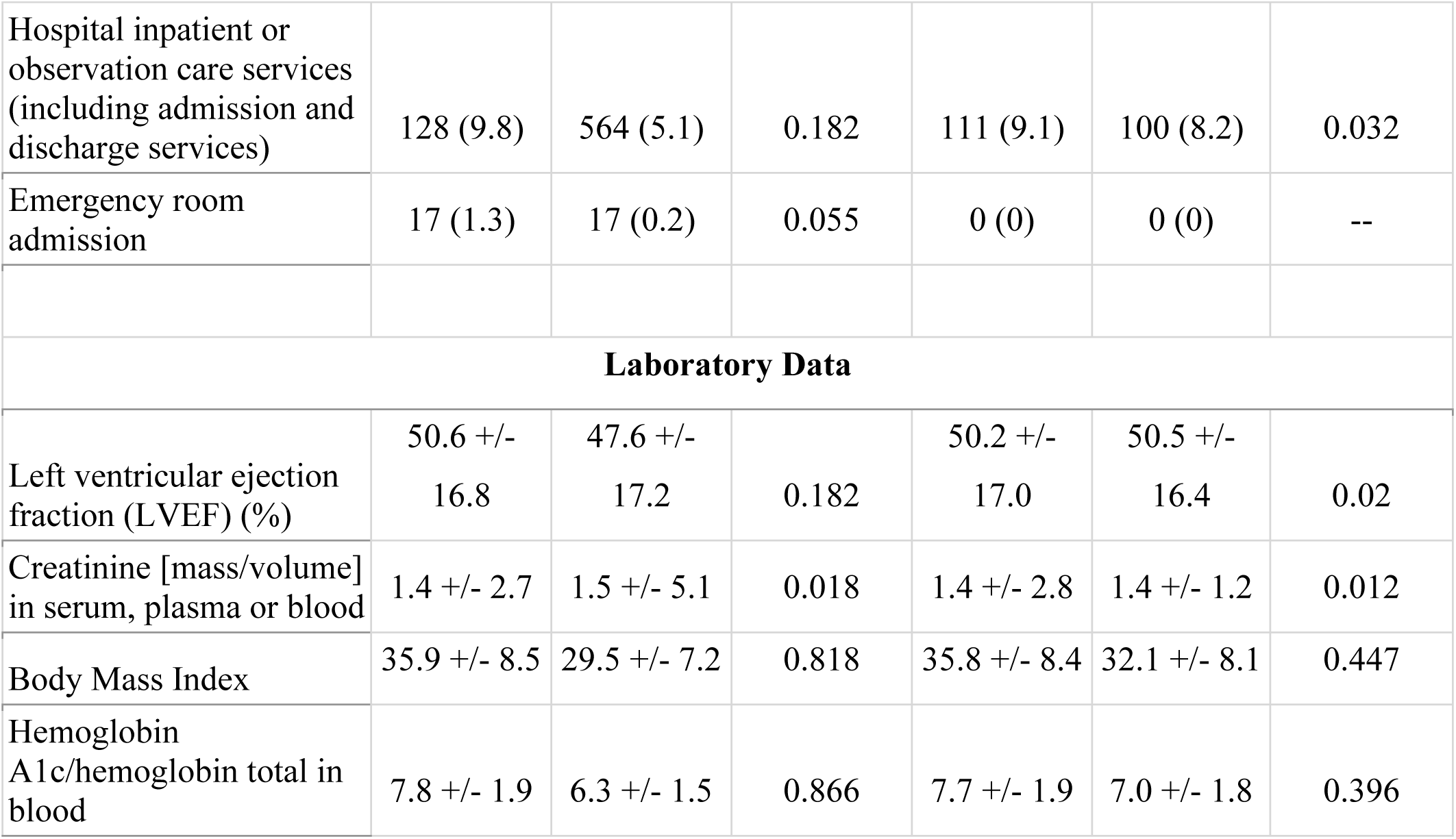
Baseline Characteristics of Study Cohort Before and After Propensity Score Matching Based on Glucagon-Like Peptide-1 (GLP-1) Analogues Treatment.

## RESULTS

### Study Population

This study identified a total of 12,410 patients who developed non-ischemic cardiomyopathy following antineoplastic therapy for cancer. Among these, 1,300 patients were on GLP-1 RA, while 11,110 were not. After propensity score matching (PSM), 1,223 patients remained in each cohort and were included in the final analysis. **Figure 1** demonstrates the patient selection process.

**Figure 1.**
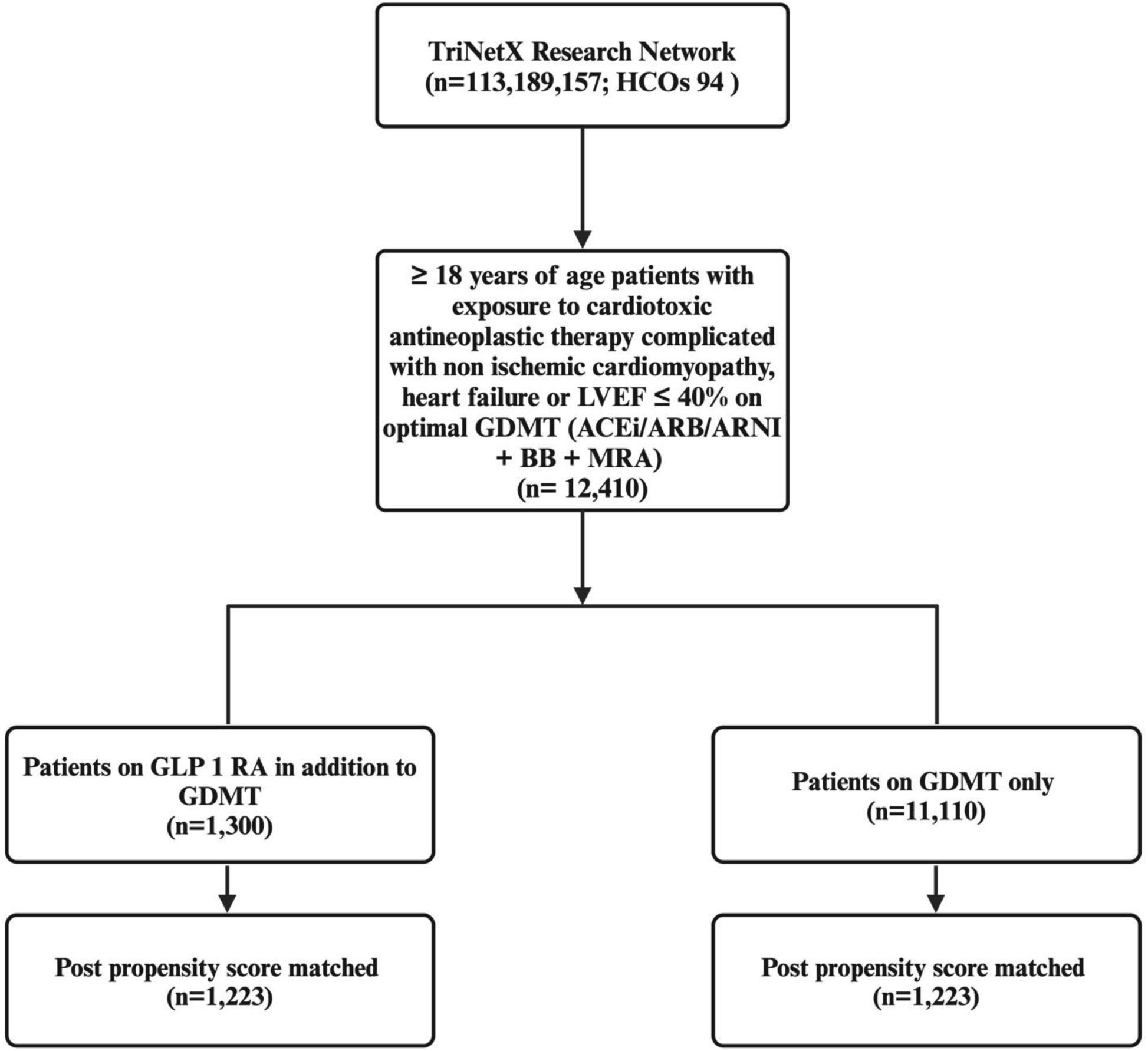
Patient Selection Process (CONSORT Diagram).

Table 1 provides a summary of the baseline characteristics of both cohorts. In the unmatched cohort, patients receiving GLP-1 RA treatment were more likely to be Hispanic or Latino (6.1% vs. 3.7%, p < 0.05) and Black or African American (22.5% vs. 13.8%, p < 0.05). Additionally, the GLP-1 RA treatment group exhibited a higher prevalence of essential (primary) hypertension, mixed hyperlipidemia, chronic kidney disease (CKD), and type 2 diabetes mellitus, but a lower prevalence of atrial fibrillation and flutter. Following PSM, the baseline characteristics of the two groups portrayed similar prevalence of the comorbid conditions described earlier.

The most common malignancies observed were neoplasms of uncertain behavior, polycythemia vera and myelodysplastic syndromes, malignant neoplasms of lymphoid, hematopoietic, and related tissue, and neoplasms of unspecified behavior. However, hematologic malignancies were most prevalent in both groups. The most frequently administered antineoplastic therapies were fluorouracil, followed by bevacizumab, cyclophosphamide, and doxorubicin. After PSM, approximately 26% of patients in each group were on bevacizumab, 14% were on doxorubicin and 4.6% were on trastuzumab.

### Study Outcomes

The primary outcome investigated was a composite of heart failure exacerbation, death, and admission to the hospital or emergency department. This outcome included a total of 681 patients in the GLP-1 RA group, compared to 843 patients in the non-GLP-1 RA group (55.7% vs. 68.9% of the cohort, respectively; OR: 0.566 [95% CI: 0.480-0.668]; p < 0.001) **(Table 2**, **Figure 2)**. Survival analysis of the primary outcome further demonstrated significant benefits of GLP-1 RA therapy (χ² = 48.274, p < 0.001, HR: 0.701 [95% CI: 0.633-0.775]) in patients with CTRCD **(Table 3)**. The secondary outcomes showed a significant decrease in mortality rate (OR: 0.471 [95% CI: 0.480-0.668]; p < 0.001), heart failure (OR: 0.661 [95% CI: 0.548-0.797]; p < 0.001), and all-cause hospital admissions (OR: 0.654 [95% CI: 0.557-0.767]; p < 0.001) among patients on GLP-1 RA **(Table 2)**. In contrast, while there was a decreasing trend for atrial fibrillation and flutter (OR: 0.932 [95% CI: 0.787-1.103]; p = 0.413), and ventricular tachycardia (OR: 0.919 [95% CI: 0.678-1.246]; p = 0.587) among GLP-1 RA patients, these results were not statistically significant **(Table 2)**. Additionally, survival analysis of the secondary outcomes further confirmed that GLP-1 RA therapy reduced the risk of cardiovascular adverse outcomes **(Table 3)**.

**Figure 2.**
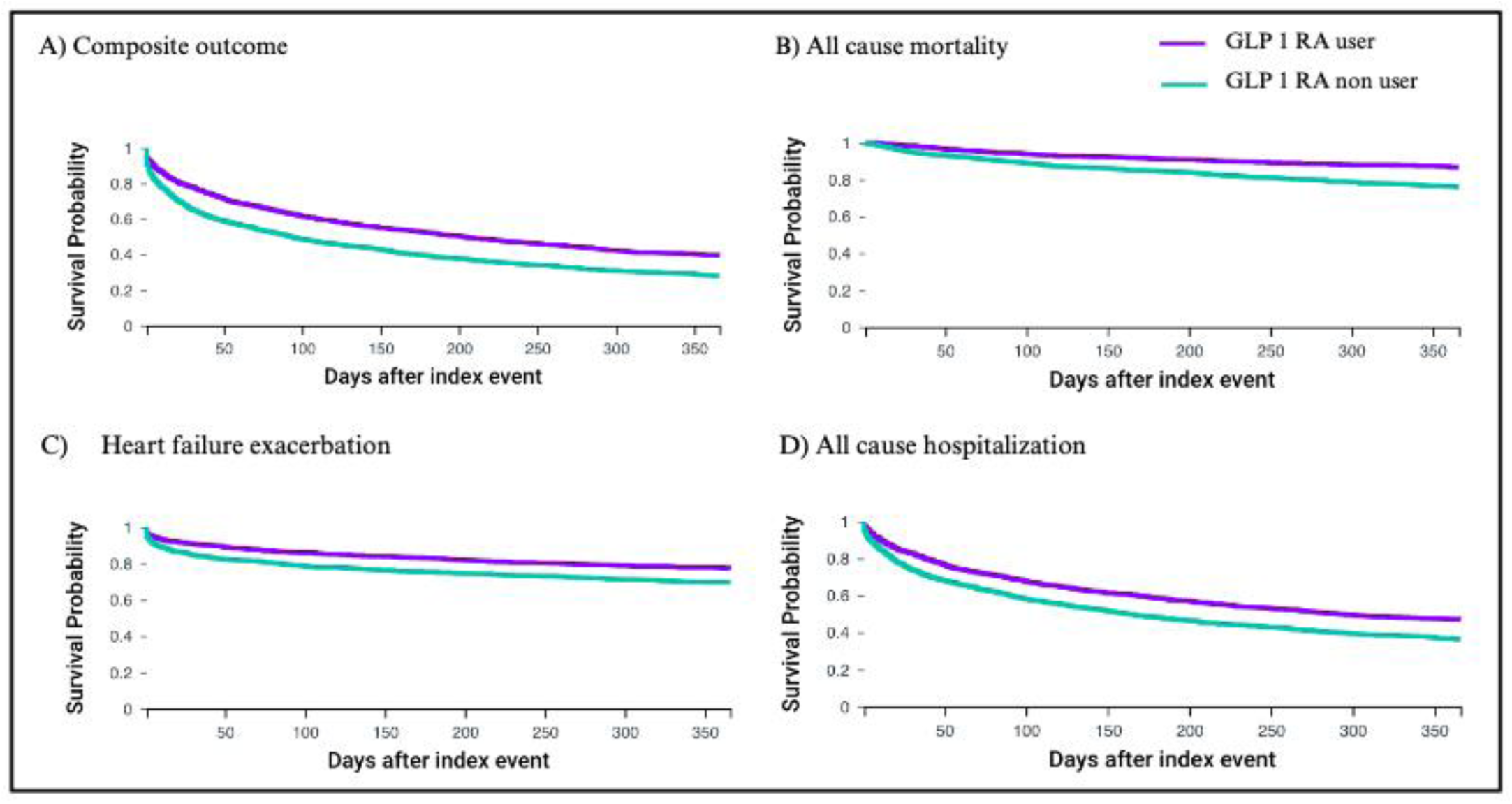
Survival probability from All-cause mortality, all-cause hospital admission or emergency room visit, Heart failure exacerbation.

**Table 2:**
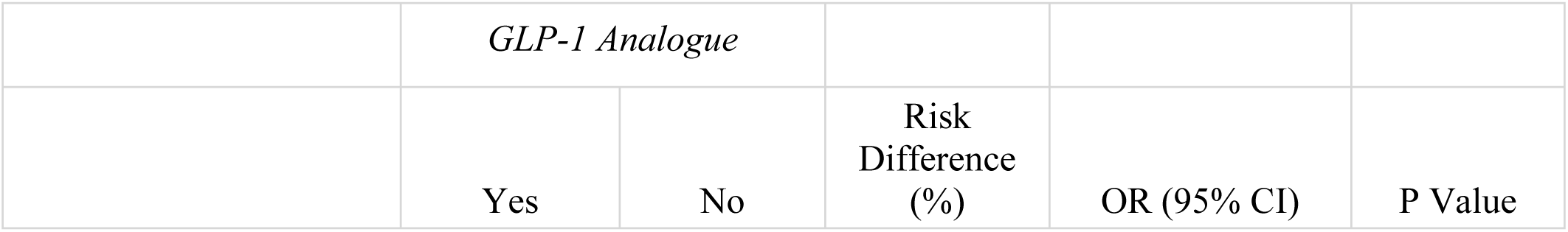
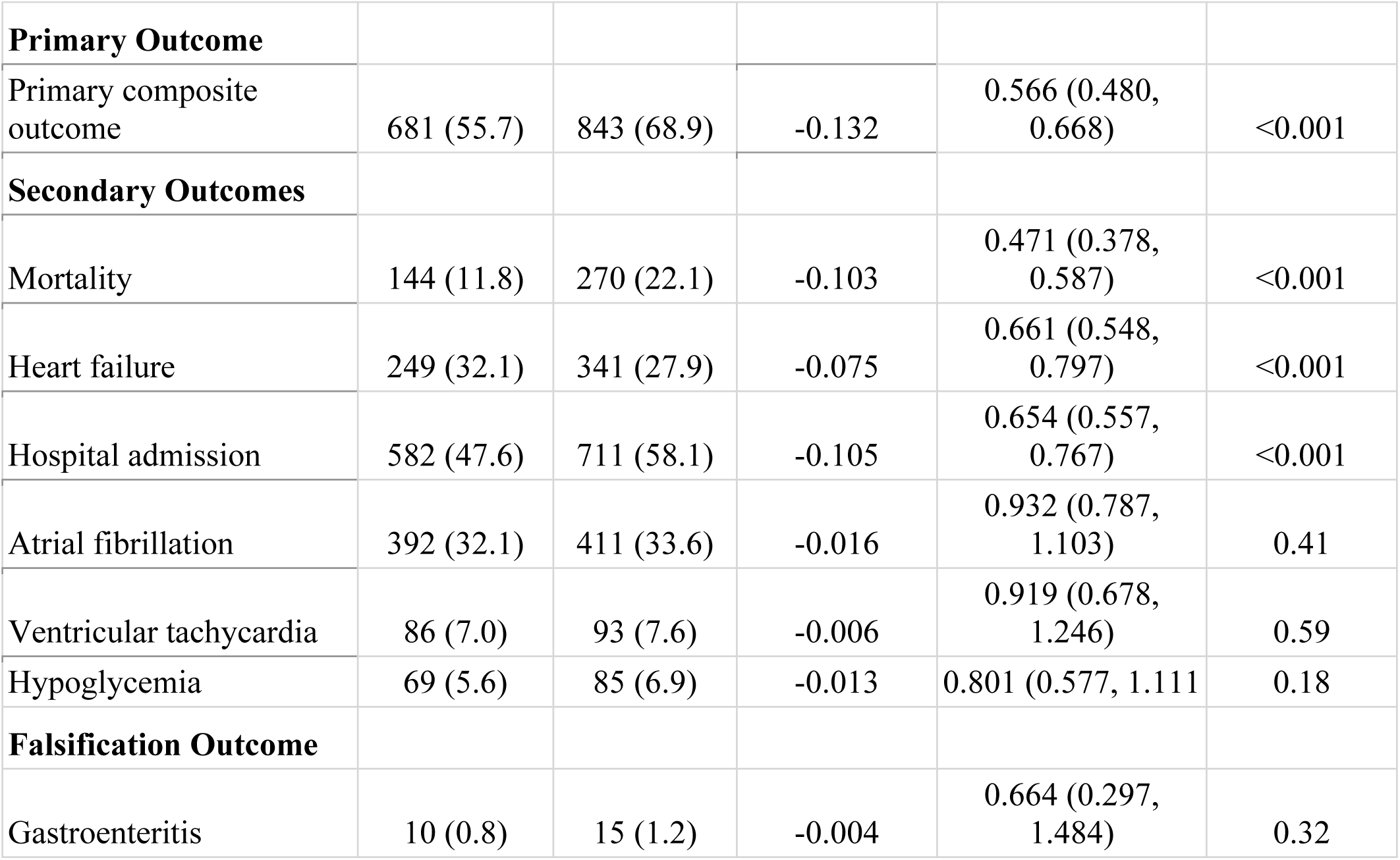
Comparison of Outcomes with and without GLP-1 Analogues in Patients with Cancer Therapy - Related Cardiac Dysfunction or Heart Failure.

**Table 3:**
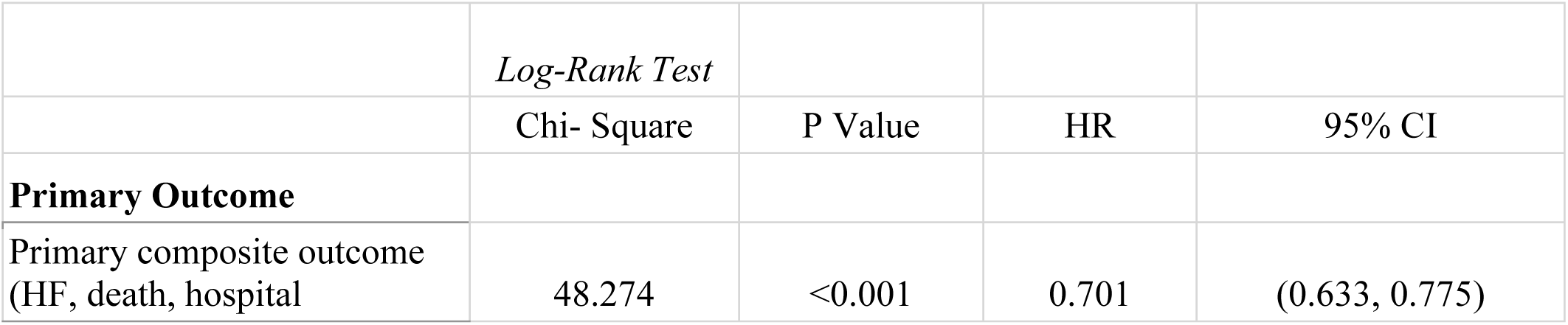
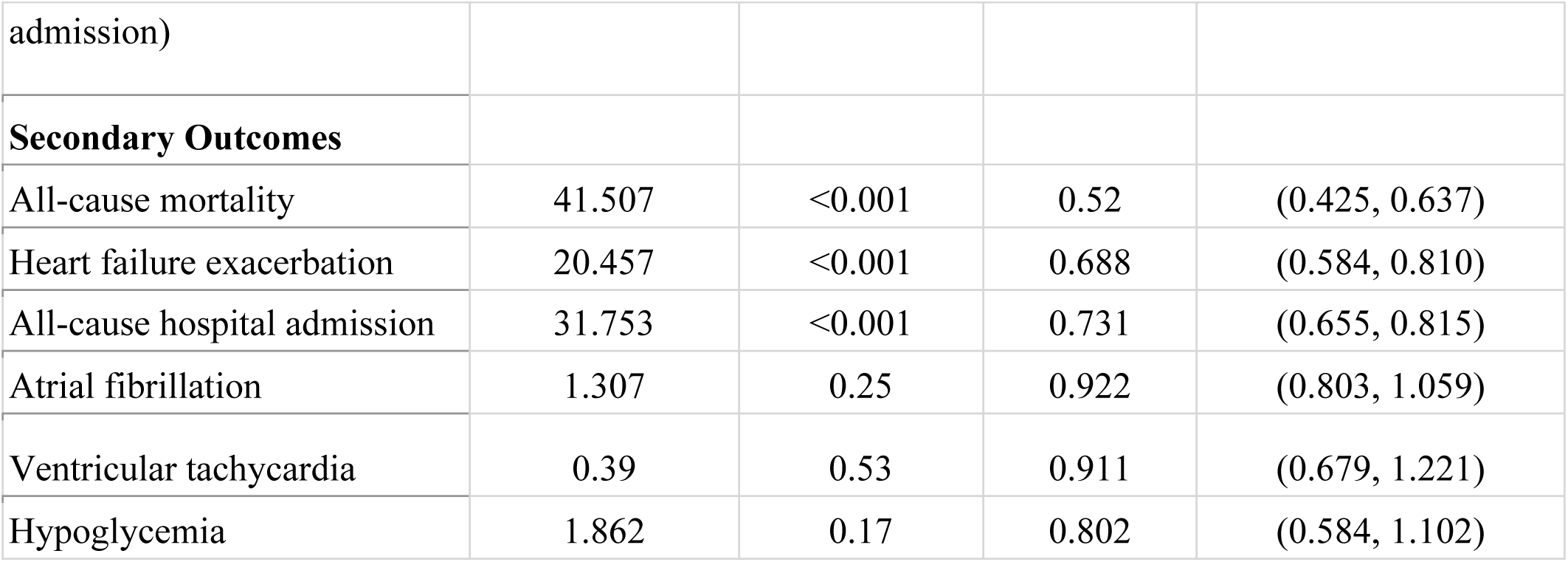
Survival Analysis of Primary and Secondary Outcome.

|  | *Log-Rank Test* |  |  |  |
| --- | --- | --- | --- | --- |
|  | Chi- Square | P Value | HR | 95% CI |
| **Primary Outcome** |  |  |  |  |
| Primary composite outcome (HF, death, hospital | 48.274 | <0.001 | 0.701 | (0.633, 0.775) |
| admission) |  |  |  |  |
| **Secondary Outcomes** |  |  |  |  |
| All-cause mortality | 41.507 | <0.001 | 0.52 | (0.425, 0.637) |
| Heart failure exacerbation | 20.457 | <0.001 | 0.688 | (0.584, 0.810) |
| All-cause hospital admission | 31.753 | <0.001 | 0.731 | (0.655, 0.815) |
| Atrial fibrillation | 1.307 | 0.25 | 0.922 | (0.803, 1.059) |
| Ventricular tachycardia | 0.39 | 0.53 | 0.911 | (0.679, 1.221) |
| Hypoglycemia | 1.862 | 0.17 | 0.802 | (0.584, 1.102) |

We checked falsification outcome to ensure the robustness of out outcomes The incidence of gastroenteritis was similar between patients with and without the GLP-1 RA, with 10 patients in the treatment group and 15 patients in the control group experiencing gastroenteritis (OR: 0.664 [95% CI: 0.297-1.484]; p = 0.315) **(Table 2)**.

## DISCUSSIONS

This is the first study to confirm the compelling benefit of GLP-1 RAs in improving cardiovascular outcomes in patients with CTRCD. There was a remarkable reduction in the individual and combined risks of death, heart failure exacerbations, and hospital admissions in CTRCD patients taking GLP-1 RAs, underscoring the protection provided by these agents. This study establishes improved overall outcomes in an often-underrepresented group of patients with the potential to transform therapeutic pathways in cancer treatment.

Our analysis reveals that amongst patients with CTRCD on GDMT, the addition of GLP1 RA led to a reduction of 52.8% in mortality and a 37.2% decrease in hospital admissions. The primary composite outcome of heart failure exacerbation, death, and hospital or emergency department admissions was also significantly lower in patients treated with GLP1. These findings align with data from LEADER, SUSTAIN-6, and SELECT clinical trials showing that GLP-1RAs in reducing overall cardiovascular events. (8,12) The rate of hospitalization was lower in our study, while the rate of hospitalization including for angina or heart failure was not significantly lower in the LEADER, SUSTAIN-6 or the SELECT trial. The distinct difference between our study and these trials was the lack of representation of patients with malignancies requiring treatment 5 years prior to inclusion. (8,12) Although our study was inclusive of patients independent of their diabetes status, a large percentage of the patients included were diabetics. (12) This is likely due to the easier access of diabetic patients to GLP1 RA. GLP-1RAs also demonstrated a numeric reduction in the incidence of atrial fibrillation (AFib) and ventricular tachycardia (VT) suggesting that compared to other cardiovascular outcomes its influence on arrhythmias may not be as profound. The effect of GLP-1 RA on arrhythmias was not studied in these major randomized control trials.

The primary cardioprotective effect of GLP-1 RA stems from its anti-oxidative and anti-inflammatory properties at the cellular level. (6,11,13) Activation of the GLP1 RA receptors in myocytes leads to downstream signaling pathways eventually increasing antioxidant proteins and pro survival kinases. (9,10) Such an effect of GLP1 RA is not limited to myocytes but is also found in vascular endothelial cells. (13) This collectively results in improved mitochondrial function, effective energy homeostasis, cellular proliferation, increased nitric oxide levels, increased angiogenesis, and decreased apoptosis (9,11,13). These cellular changes are histologically translated to a dose-dependent significant reduction in degenerative myocardial and inflammatory cells observed in murine myocardial tissues pre-treated with semaglutide, exposed to doxorubicin (9). Biochemically this effect appears as a decreased level of CK-MB and cardiac troponin which are often elevated in patients with CTRCD. (9, 11) Together these effects underline the substantial cardioprotective properties of GLP-1RAs, highlighting their potential as a valuable therapeutic option for patients on anti-cancer treatments without attenuating their anti-cancer properties (10).

Although GLP-1RAs are known for their efficacy in managing type 2 diabetes mellitus and their favorable safety profile in heart failure patients, they are not exempt from adverse events. Transient nausea and vomiting are the most common adverse reactions with GLP-1RAs (14). More serious concerns are of these drugs with pancreatitis, pancreatic cancer, and thyroid cancer, however, at present there is no conclusive evidence to support it (15,16). The number of patients with these malignancies was extremely limited in our data to evaluate for any realistic benefit in them.

There are notable limitations to our study. Firstly, the observational nature of the research means causality cannot be definitively established. Although propensity score matching was employed to control confounding factors, residual confounding may still influence the results. Additionally, the study’s reliance on electronic health records limits detailed data collection on patient adherence to GLP-1RA therapy and other non-pharmacological interventions. Also, chemotherapy regimens usually consist of a combination of multiple anti-neoplastic drugs. The study was unable to evaluate the benefit of GLP1 RA for chemotherapy regimens given the inability to query the database for them. Future randomized controlled trials will help validate these findings and explore the underlying mechanisms of GLP-1 RA in patients with CTRCD.

## LIMITATIONS

This study has several limitations to be considered when interpreting the results. First, the results rely heavily on the clinicians’ accurate documentation. Secondly, the database lacks a few granular information, including the combination of anti-neoplastic therapies used in each patient. Thirdly, there is limited diversity in the demographic groups. Certain demographic groups, such as underrepresented minorities, may not have been adequately represented in the dataset. Lastly, the follow-up period for patients was relatively short, restricted to one year. This limited time frame makes it difficult to assess long-term outcomes.

## CONCLUSIONS

GLP-1RAs represent a promising therapeutic option for patients with CTRCD, offering substantial benefits in reducing mortality, heart failure exacerbations, and hospital admissions. The overall cardioprotective benefits of GLP-1RAs support their integration into treatment strategies for cancer survivors with cardiac dysfunction.

## Data Availability

Data can be obtained from TriNet X research database. Supplemental file includes the diagnostic codes for the data.

## Abbreviations

CTRCD: Cancer therapy-related cardiac dysfunction
GLP-1 RAs: Glucagon-like peptide-1 receptor agonists
GDMT: Guideline-directed medical therapy
LVEF: Left ventricular ejection fraction
HF: Heart failure
T2DM: Type 2 diabetes mellitus
ACEI: Angiotensin-converting enzyme inhibitors
ARB: Angiotensin receptor blockers
ARNI: Angiotensin receptor/neprilysin inhibitor
BB: Beta blocker
MRA: Mineralocorticoid receptor antagonist
CABG: Coronary artery bypass grafting
PCI: Percutaneous coronary intervention

---

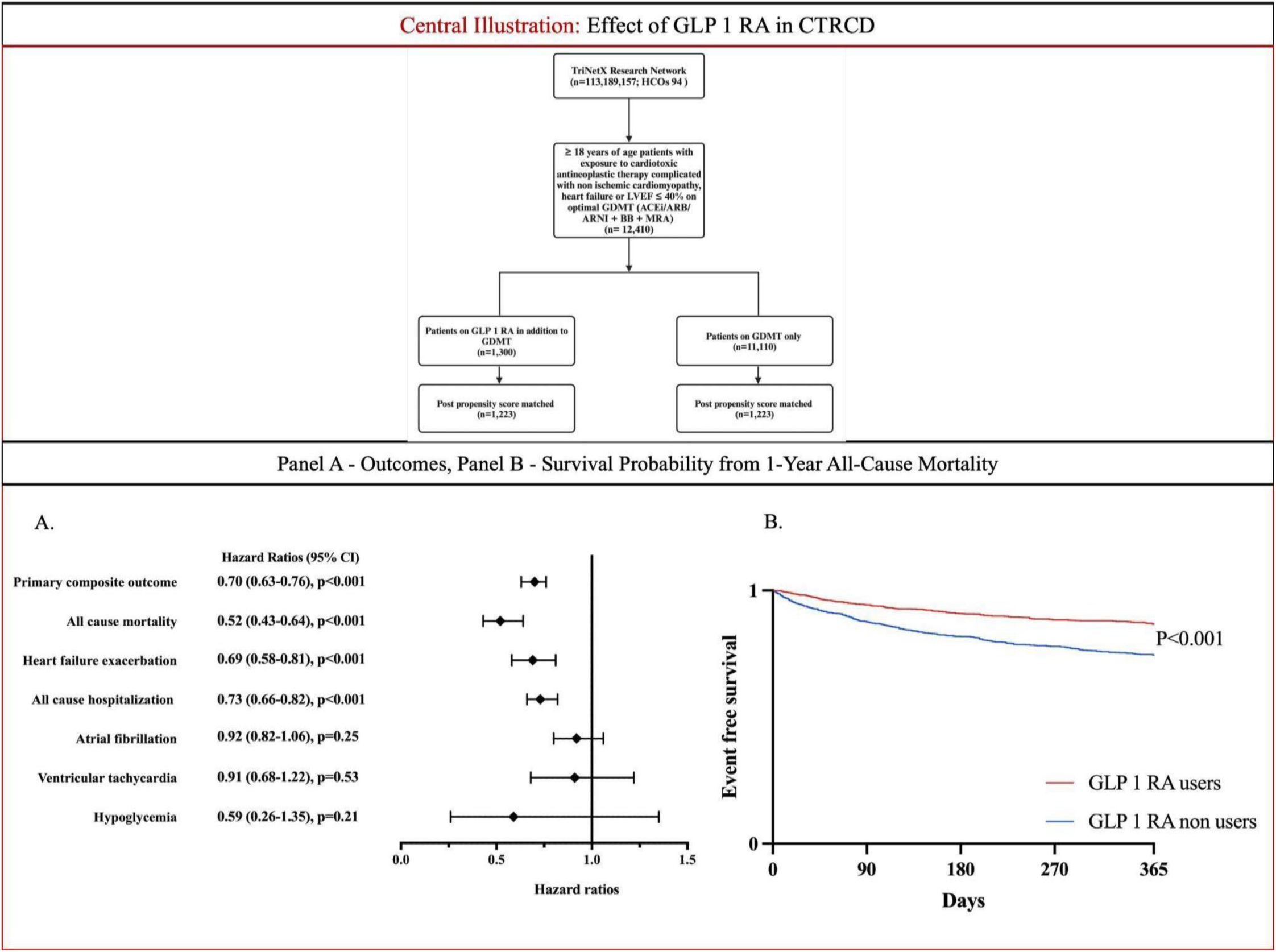
**Central illustration**

## Notes

### Competing Interest Statement

The authors have declared no competing interest.

### Funding Statement

This research received no specific grant from funding agencies in the public, commercial, or not-for-profit sectors.

### Author Declarations

Data is deidentified, and Institutional Review Board approval is not required. Analysis done in TrINetX platform.

